# Children develop Immunity to cryptosporidiosis in a high transmission intensity area

**DOI:** 10.1101/2023.06.28.23292000

**Authors:** William A. O. Petri, Biplop Hossain, Mamun Kabir, Hannah H So, G. Brett Moreau, Uma Nayak, Jennie Z Ma, Zannthan Noor, ASG Faruque, Masud Alum, Rashidul Haque, William A Petri, Carol A Gilchrist

## Abstract

**Background:** *Cryptosporidium* is one of the top causes of diarrhea in Bangladesh infants. *Cryptosporidium* infections lead to the production of antibody immune responses, which were associated with a decrease in parasite burden and decreased disease severity in subsequent infections.

**Methods:** We conducted a longitudinal study of cryptosporidiosis from birth to five years of age in an urban slum of Dhaka Bangladesh. We then retrospectively tested the concentration of anti-Cryptosporidium Cp17 or Cp23 IgA in surveillance stool samples collected from 54 children during their first 3 years of life by enzyme-linked immunosorbent assay (ELISA). We also assessed the concentration of both IgA and IgG antibodies specific to Cryptosporidium Cp17 and Cp23 in the concentration of anti-Cryptosporidium Cp17 or Cp23 IgA and IgG antibodies in the children’s plasma (1-5 years).

**Results:** The seroprevalence of both anti-Cp23 and Cp17 antibodies was high at ≤ one year of age and reflected the exposure of these children in this community to cryptosporidiosis. In Bangladesh, the prevalence of cryptosporidiosis is high during the rainy season (June to October) but decreases during the dry season. In younger infants’ plasma anti-Cp17 and Cp23 IgG and anti-Cp17 IgA levels were markedly increased during the rainy season in line with the higher initial exposure to the parasite at this time. Both anti-Cp17, anti-Cp23 fecal IgA and the parasite burden declined during repeat infections.

**Conclusions:** We found that anti-Cryptosporidium plasma and fecal antibody levels in children could contribute to the decrease in new infections in this study population.

## Introduction

*Cryptosporidium* is one of the top causes of diarrhea and growth faltering in Bangladeshi infants. We previously carried out a natural history study of cryptosporidiosis, tracking infants’ infections from birth until they reached 3 years of age (National Clinical Trial Identifier: NCT02764918) (Gilchrist et al. 2018; Steiner et al. 2018, 2020, 2021). These infants were inhabitants of lower-income neighborhoods within Dhaka, Bangladesh. The intensive surveillance undertaken during years 1-3 of their lives, combined with the high exposure to cryptosporidiosis in this population, allowed us to analyze the impact of repeated infections (sub-clinical and symptomatic) on Bangladeshi child health (Kabir et al., 2021). The children’s immunity to repeated infection was lacking, as most children were reinfected before they turned 3 years of age. In this paper, we present the data gained by following the children up to 5 years of age. Diarrheal cryptosporidiosis decreased rapidly during this period. To investigate the potential role of mucosal and systemic immunity, samples from a subgroup of 54 children previously infected before 1 year of age were examined. Antibodies against the Cryptosporidium antigens Cp23 and Cp17 were measured in both surveillance fecal samples collected between 0-3 years of age (IgA only) and in the plasma samples collected between 1-5 years (both IgA and IgG).

## Results

### Cryptosporidiosis frequency

Five hundred children were enrolled within the first week of birth, and of these 286 completed five full years of observation (Figure 1). Stool samples were collected during diarrheal episodes (0-5 years). Although diarrheal samples continued to be collected from all of the children enrolled in the 5 years of the study, in years 3-5 the monthly surveillance was only continued for children in Sub-Group 1 with surveillance samples being collected at only 6-month intervals in Sub-Group 2 (Figure 1). In our study cohort, the peak of Cryptosporidium associated diarrhea occurred at ∼ 2 years of age and thereafter fell rapidly with oldest *Cryptosporidium* positive diarrheal sample being collected from a child 4.3 years in age. The data on Cryptosporidium-associated diarrhea prevalence <4 years in age (Figure 2) was therefore unlikely to be affected by either the change in surveillance sample collection or by the hiatus in stool collection between April 2020 to August 2020 due to the COVID-19 pandemic (Supplemental Table 1) which depressed the number of surveillance stools collected in children between 4.5-5.0 years in age (Figure 3A). To confirm that the decrease in surveillance did not inflate the number of cryptosporidium infections associated with diarrhea the data from the different collection protocols (Sub-Group 1 and Sub-Group 2) were analyzed separately and no difference in infection profile was observed (Supplemental Figure 1). The incidence of diarrhea-associated cryptosporidiosis therefore in our study cohort appeared to decrease rapidly over the 3-4 year observation period (Figure 2).

**Figure 1.**
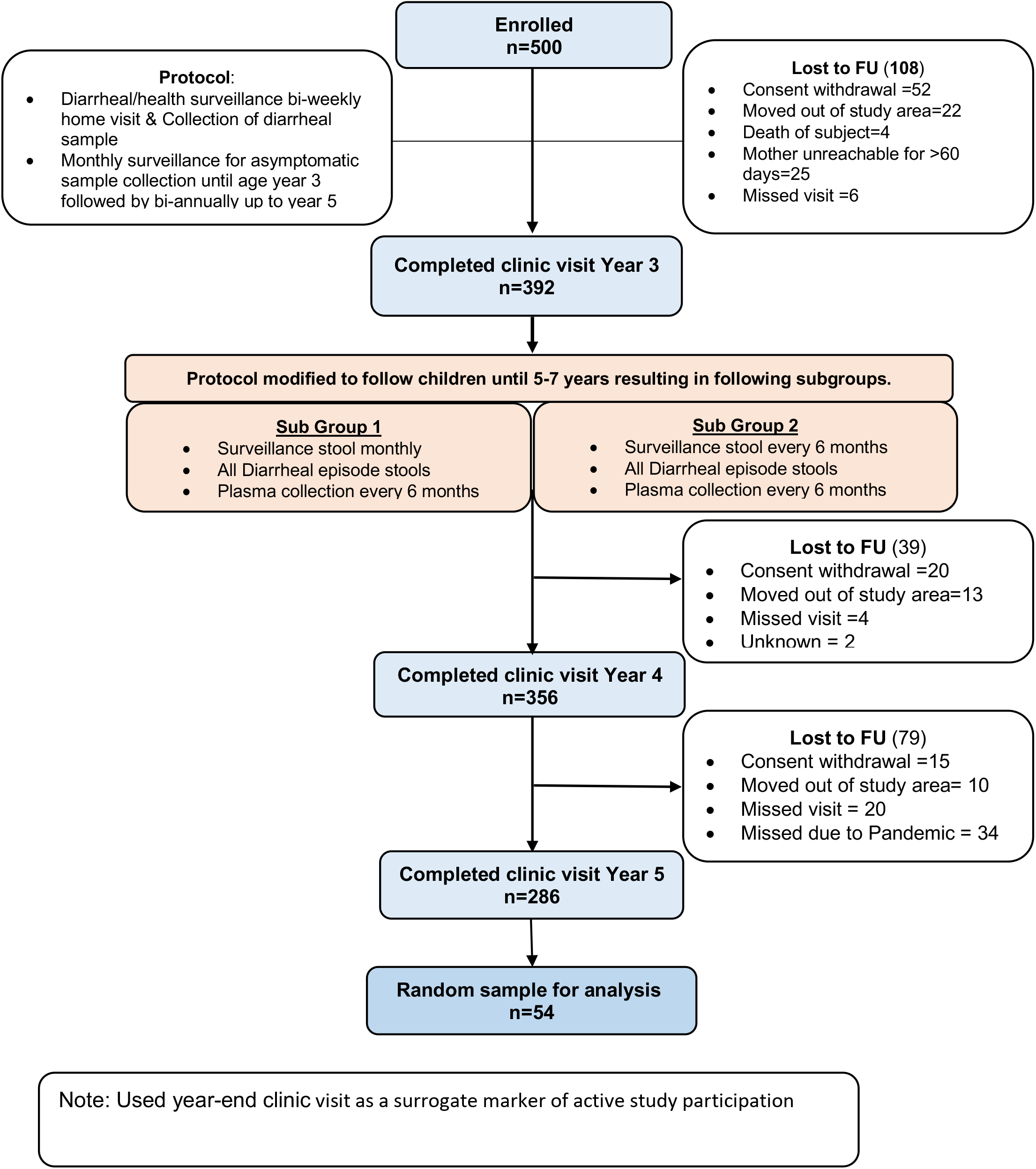
Flow diagram of Birth COHORT Study.

**Figure 2.**
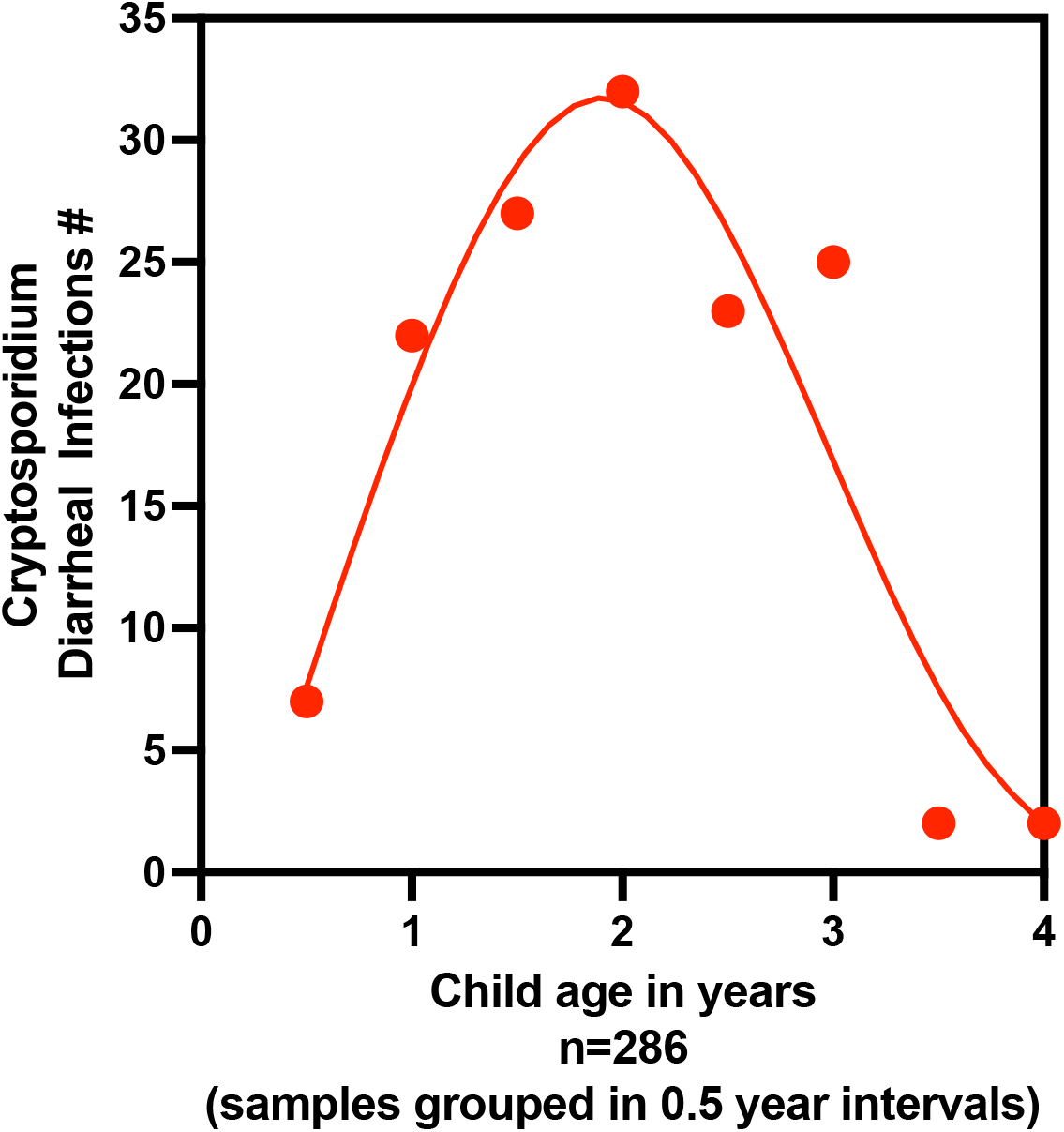
Incidence of Diarrheal Cryptosporidiosis in the Mirpur cohort. Data shown is only from the 286 children who remained in the study for the entire 5 years; Diarrheal cryptosporidiosis phenotype is defined as an infection in which symptoms were coincident with the qPCR detection of Cryptosporidium in fecal DNA. Positive samples were classified as a separate infection only if occurring greater than 65 days after the preceding positive sample. Infections occurring within each 6-month interval were grouped and the number shown on the Y axis. X axis indicates child age in days. Although cryptosporidiosis was known to decrease in frequency diarrheal stool collection and analysis took place > 3 years of age although there was a decrease in surveillance. All of the children in the group of 286 children shown in this graph had reached 4 years of age prior to the COVID-19 pandemic-related hiatus in surveillance sample collection (Supplemental Table 1).

**Figure 3:**
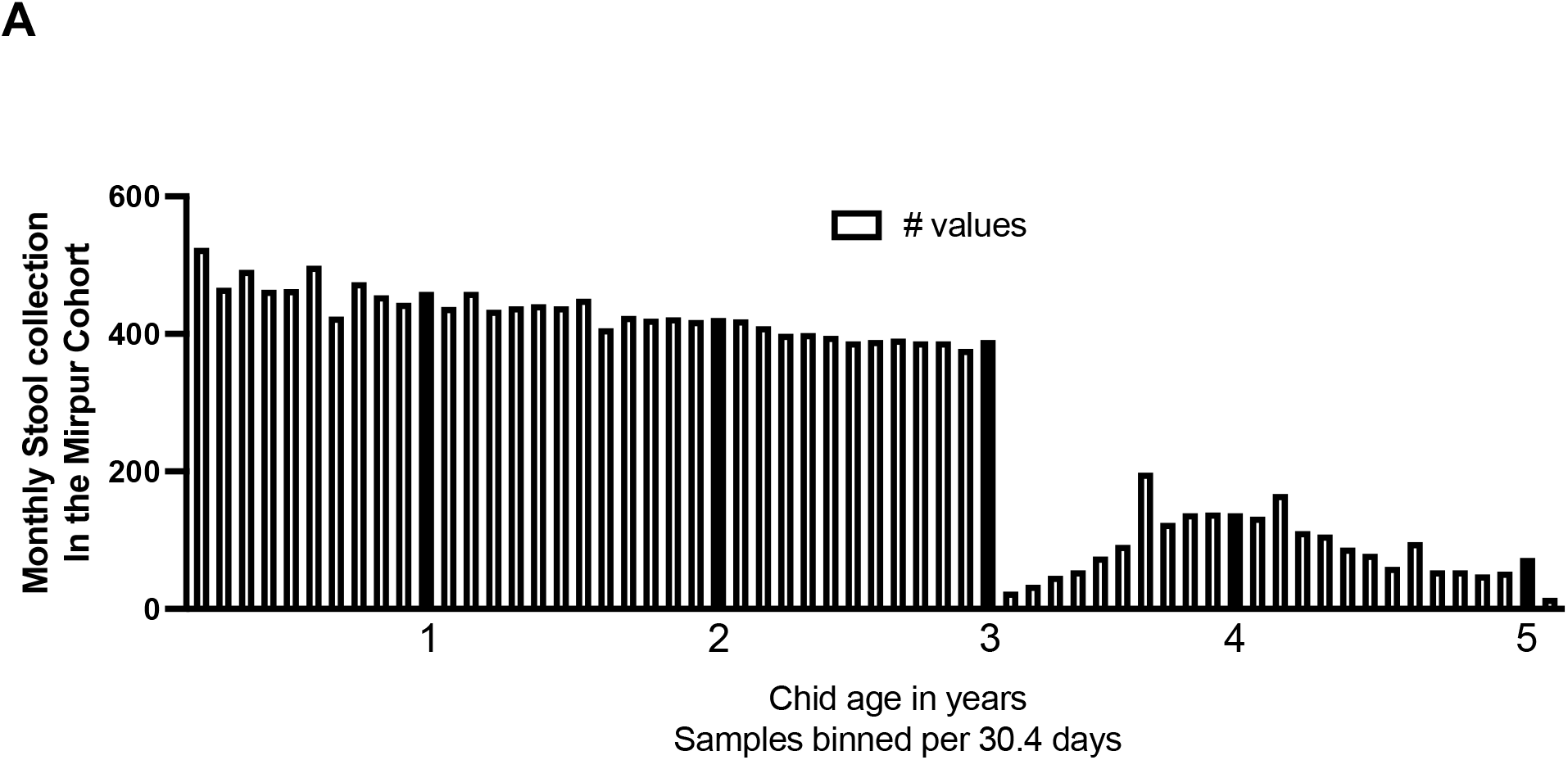
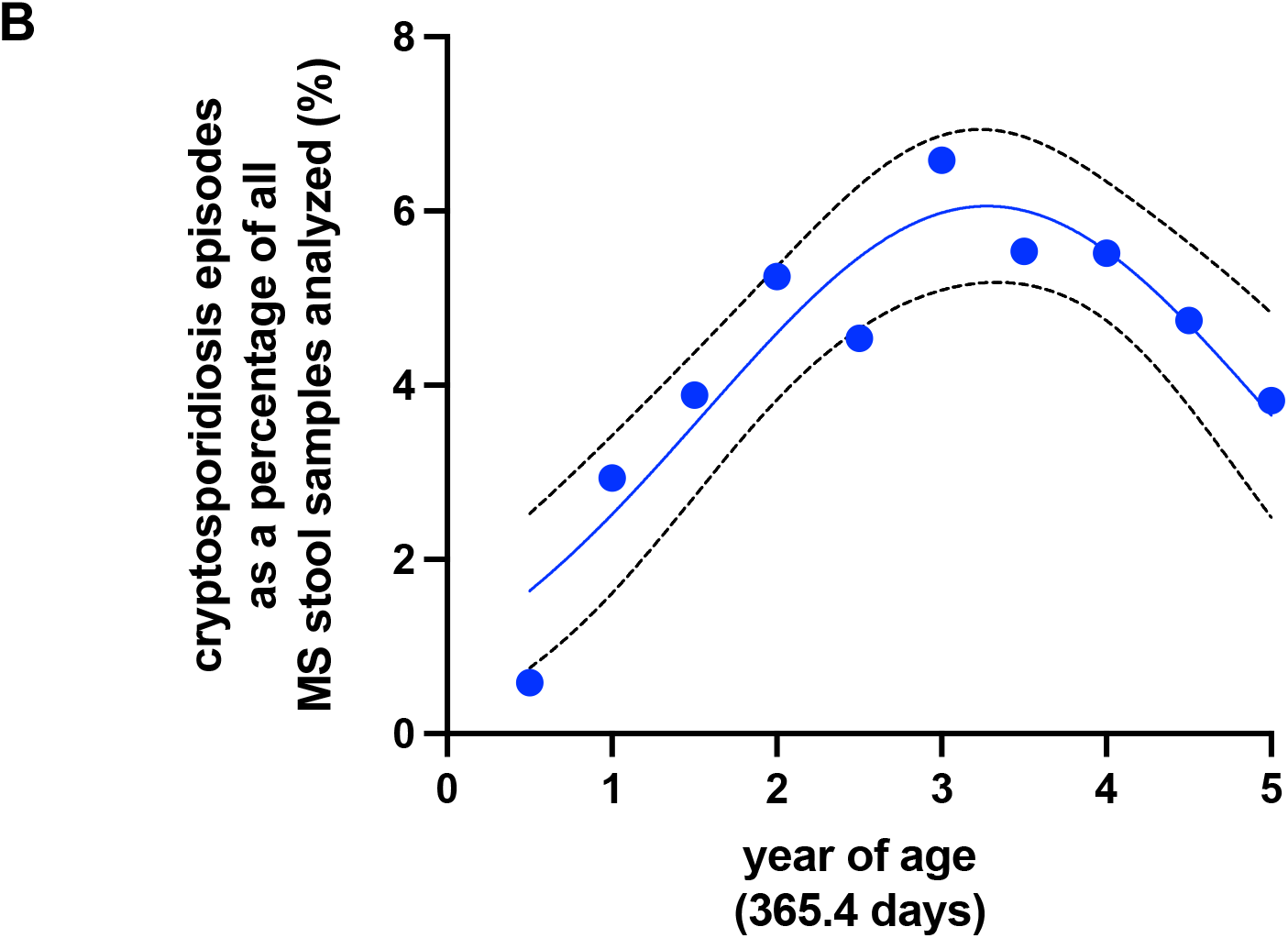
Decline in the frequency of sub-clinical cryptosporidiosis in the Mirpur cohort. A) Sub-clinical cryptosporidiosis phenotype is defined as an infection in which no symptoms were observed at the time of qPCR detection of Cryptosporidium in fecal DNA. Stool surveillance was decreased after the children reached 3 years of age with monthly stool samples being collected from only a third of the infants, all other collections occurred bi-annually. Y axis indicates the number of fecal samples collected during each 30.4-day window (year end is indicated by the filled bar). The graph clearly shows the transition at year 3 from continuing monthly surveillance to a proportion of the cohort switching to bi-annual cross-sectional surveillance B) Sub-clinical cryptosporidiosis is expressed as a percentage of the number of samples collected to account for the differences in collection frequency. Y-axis indicates the percentage of infections binned into 6-month intervals as shown on the X axis which indicates the child’s age in years. The dotted lines indicate the 95% CI of the non-linear curve fit line.

To compensate for the difference in the collection of surveillance stools the analysis of the sub-clinical cryptosporidiosis disease was examined as a percentage of the number of surveillance stools collected within the previous 6-month interval (Figure 3A). The number of sub-clinical cryptosporidiosis infections remained high during year 4 (Figure 3B). An additional follow-up period is needed to see this continues and if, as occurs in young infants, sub-clinical cryptosporidiosis in older children accentuates the effects of food insecurity and results in growth faltering (Donowitz et al. 2016; Korpe et al. 2016; Mondal et al. 2012; Schnee et al. 2018; Steiner et al. 2018).

### Anti-Cryptosporidium Cp23 and Cp17 IgA and IgG Profiles

The 54 children selected for additional analysis (number of plasma samples analyzed n=378 and fecal samples n=1836) were randomly selected from among the children who had at least one cryptosporidium infection prior to one year in age (Figure 4A). These children had a median of 3 *Cryptosporidium* infections over the 5 years tracked, with a minimum of 1 infection and a maximum of 6 infections. (Figure 4A).

**Figure 4.**
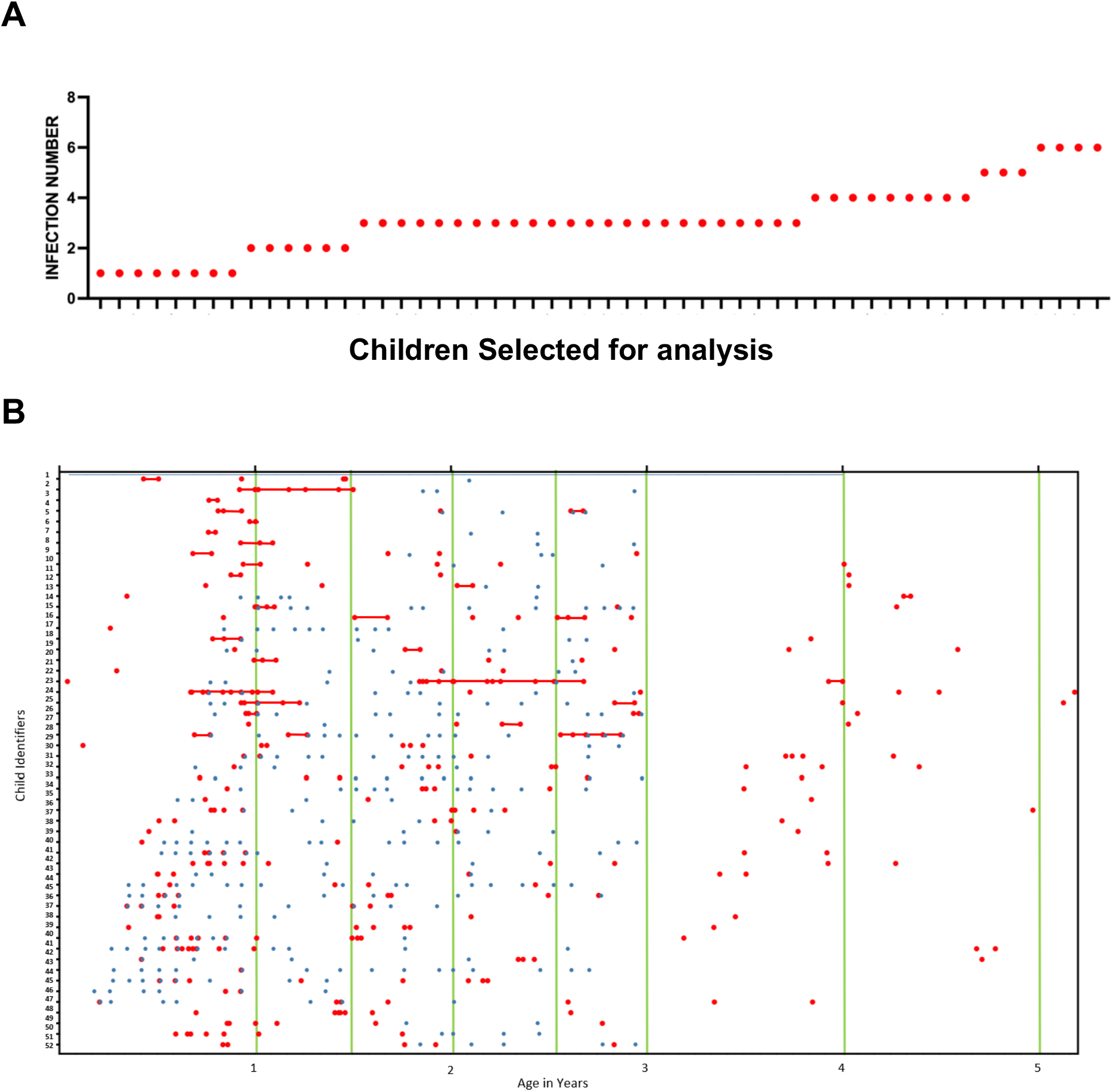
Infections and samples collected in a subset of infants selected for additional analysis A) Infections Child Within 5 Years of Birth Y axis indicates the number of infections X axis the individual infants selected for this analysis B) On the X-axis is the child’s age in years. On the Y-axis is the Child’s unique ID. Green lines represent Plasma Collection time points, the red dots represent stool samples that tested positive for Crypto, and the blue dots represent stool samples that tested positive for both Cp17 and Cp23 IgA & IgG.

One year post plasma collection, a significant proportion of pediatric subjects were serologically positive for CP17 IgG (74%) and CP17 IgA (65.8%) Cp23 IgG (73%) Cp23 IgA (43%) (Table 1).

**Table 1.** Prevalence of seropositive samples. This table shows time post birth of the plasma collection, and the percent of children with a positive signal via ELISA for Cp17 IgG and IgA antibody % positives is shown in the brackets.

| Age Range<br>(days) | Anti-Cp17 IgG |  | Anti-Cp17 IgA |  | Anti-Cp23 IgG |  | Anti-Cp23 IgA |  |
| --- | --- | --- | --- | --- | --- | --- | --- | --- |
|  | positive | negative | positive | negative | positive | negative | positive | negative |
| [182-365]<br>1 years | 22 (73%) | 8 | 19 (63%) | 11 | 22 (73%) | 8 | 13 (43%) | 17 |
| [365-548]<br>1.5 year old | 35 (67%) | 17 | 32 (61%) | 20 | 40 (77%) | 12 | 24 (46%) | 28 |
| [548-730]<br>2 years old | 40 (68%) | 19 | 32 (54%) | 27 | 52 (88%) | 7 | 24 (41%) | 35 |
| [730-913]<br>2.5 years old | 28 (85%) | 14 | 24 (57%) | 18 | 35 (83%) | 7 | 17 (40%) | 25 |
| [913-1096]<br>3 years old | 46 (86%) | 8 | 33 (61%) | 21 | 48 (89%) | 6 | 26 (48%) | 28 |
| [1096-1278]<br>3.5 years old | 18 (78%) | 3 | 15 (71%) | 6 | 19 (90%) | 2 | 9 (43%) | 12 |
| [1278-1461]<br>4 years old | 14 (77%) | 4 | 11 (61%) | 7 | 16 (89%) | 2 | 10 (56%) | 8 |
| [1461-1644]<br>4.5 years old | 17 (61%) | 11 | 17 (61%) | 11 | 24 (86%) | 4 | 14 (50%) | 14 |
| [1644-1827]<br>5 years old | 8 (47%) | 9 | 7 (41%) | 10 | 13 (76%) | 4 | 8 (47%) | 9 |

Cryptosporidiosis in Bangladesh occurs more frequently in the rainy season (June -October) (Steiner et al. 2018). We therefore examined our data not only by child age and antibody isotype but also depending on whether the samples were collected during the rainy or dry seasons. Early in life, the plasma IgG values and anti-Cp17 IgA values were markedly higher in younger children during the rainy season possibly reflecting an increased exposure to the parasite (Figure 5A-C). Interestingly, thereafter the antibody levels declined. At the five-year mark there was also a corresponding modest non-significant decrease in the number of seropositive samples possibly reflecting the sensitivity limit of the ELISA assays (delineated in Table 1).

**Figure 5.**
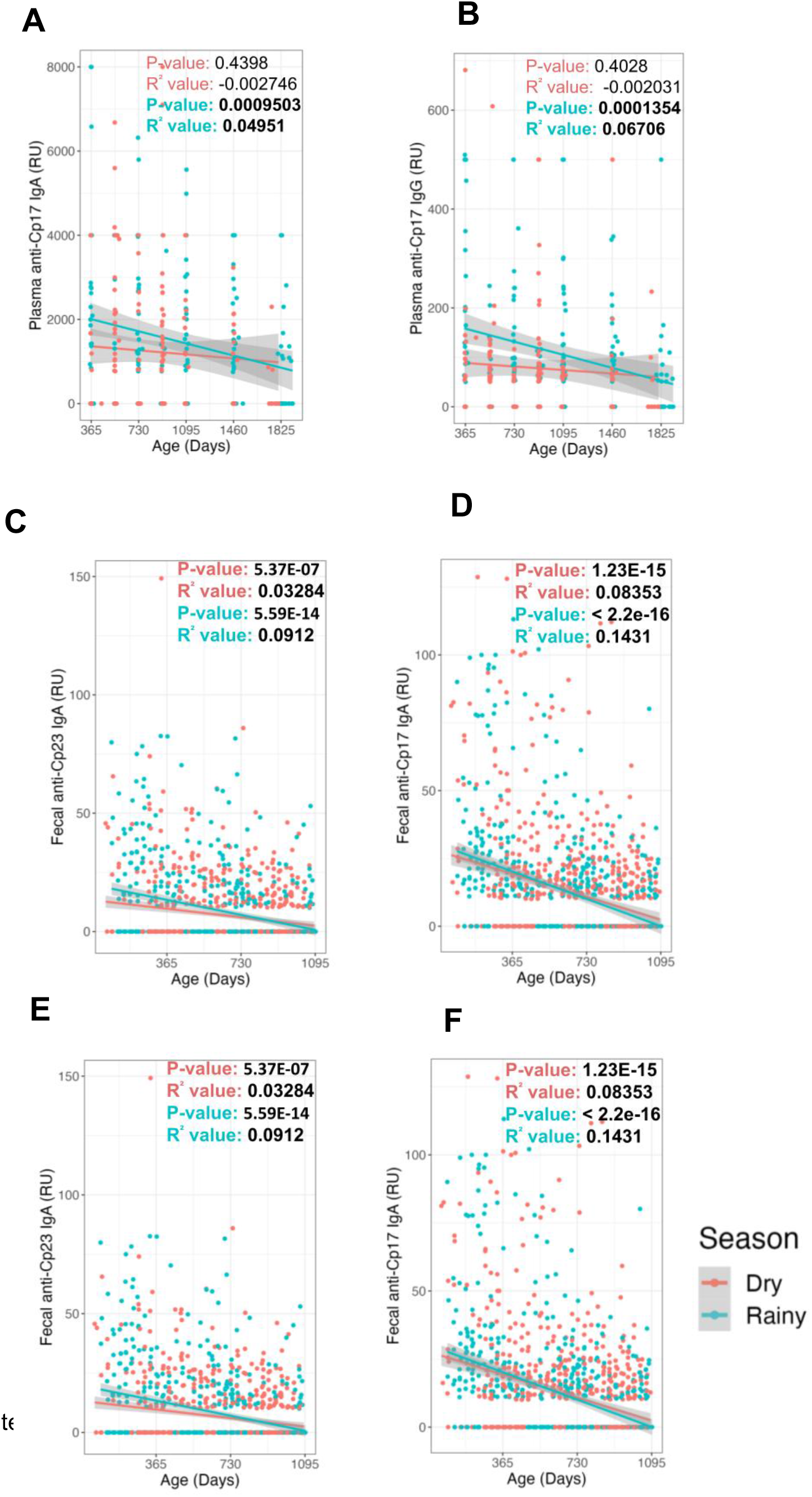
Analysis of the seasonal IgG (A & B) and IgA antibodies (C-F) to the Cp17 (A,C,E) and Cp23 (B,D,F) antigens in plasma (A-D) and fecal material (E&F). Relative antibody levels are shown on the Y-axis with the children’s age in days shown on the X-axis. Blue symbols are samples collected during the Bangladesh rainy season (June to October); red symbols dry season. Linear regression P-values and R^2^ values are shown for IgG and IgA, as well as a line (colored per season) and confidence intervals (gray bands) fit to each.

Fecal IgA values irrespective of seasonality were higher early in life (Figure 5 E, F) perhaps reflecting the higher levels of exposure of the gut immune system to this parasite. We examined the relationship between stools collected that tested positive for Cryptosporidium, and stools collected that tested positive for both fecal Cp17 and Cp23. Within the cohort of 54 pediatric subjects sampled across multiple time points, the fecal samples marked with red dots indicated a positive Cryptosporidium infection diagnosis via qPCR. In contrast, the blue dots corresponded to fecal samples that tested positive for both CP17 and CP23 IgA (Figure 4B).

To examine the impact of repeated infections on antibody levels we restricted our analysis to samples where the extracted fecal DNA was qPCR positive for the Cryptosporidium parasite and the sample was available for analysis by our in-house ELISA assays for fecal anti-Cp17 and Cp23 IgA values (Figure 6). The *Cryptosporidium* infection number was determined using the entire study dataset. The parasite burden was decreased in repeat infections as expected (Kabir et al. 2021)(Figure 6A). We found that fecal anti-Cp17 and Cp23 IgA levels were also significantly decreased (Figure 6B & C).

**Figure 6.**
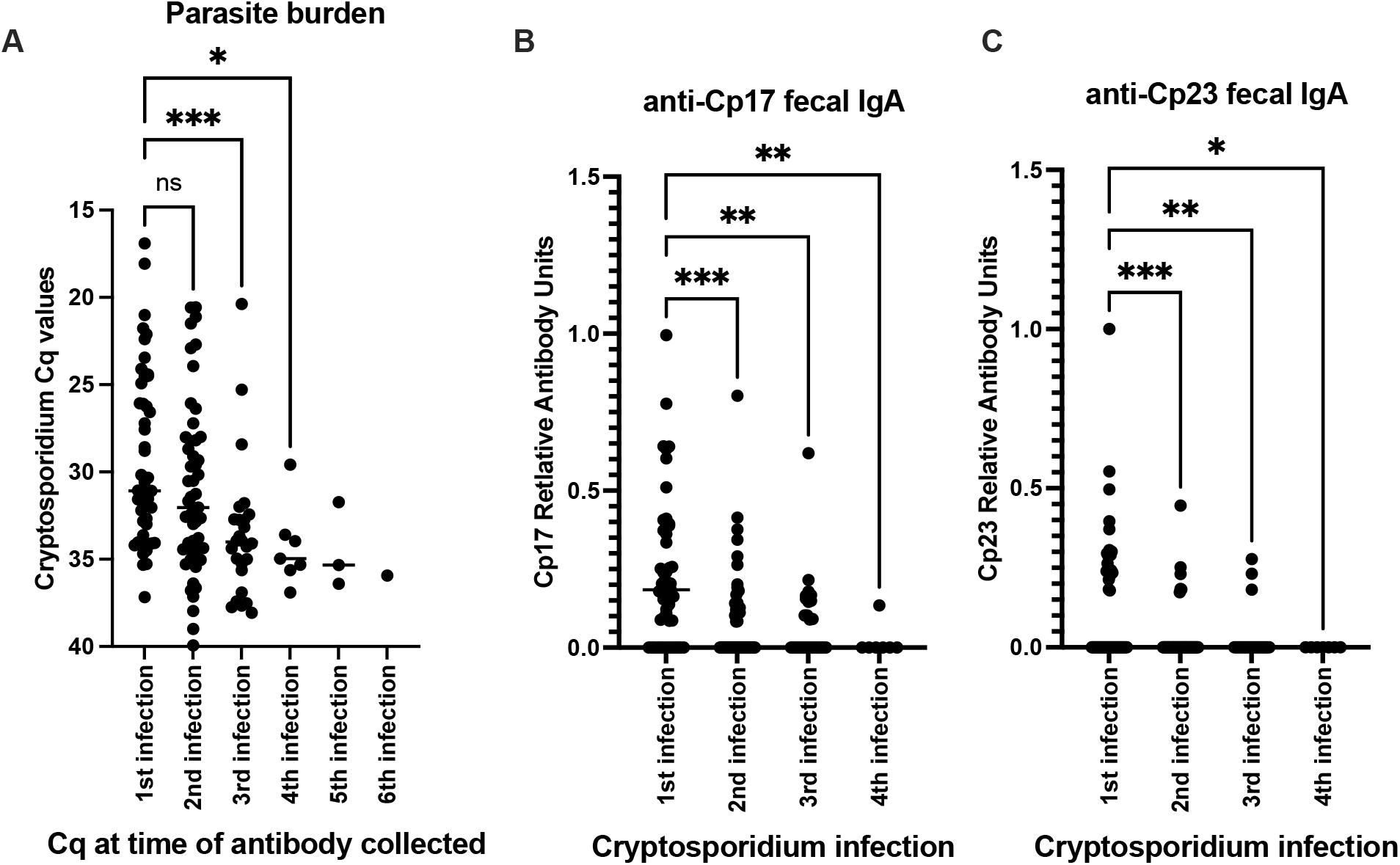
Parasite burden and anti-Cp17 and Cp23 fecal IgA decline in repeat infections. Analysis was restricted to samples in which both fecal IgA was able to be measured and the sample had a Cryptosporidium positive Cq value. X values: The number of repeat *Cryptosporidium* infections per child was defined using the entire data set as previously defined (Kabir et al 2021). A) In line with previous observations the parasite burden was decreased during repeat infections. Fecal IgA B) anti-Cp17 and C) anti-Cp23 also declined during repeat infections. (*) p value is less than 0.05, (**) p value less than 0.01, (***) p value is less than 0.00. Analysis was restricted to paired 1^st^ and 2^nd^ infections (n=19) and repeated using the mixed-effect model (Supplemental Figure 2) p=0.0041

## Discussion

Previous studies have shown that the *Cryptosporidium* antigens Cp17 and CP23 in blood plasma are associated with protection from reinfection in children through 3 years of life (Kabir et al. 2021).

The key finding of this paper is that the number of diarrheal cryptosporidium infections declined as the children grew in age and in younger children the levels of anti-*Cryptosporidium* antibodies to Cp17 and Cp23 are higher in fecal material earlier in life when first infections were common.

This observation suggests that the systemic antibody elevation due to new first infections was short-lived (Crompton et al. 2010). However, in the fecal material collected from the younger children, IgA antibodies were high in samples collected during both rainy and dry seasons possibly reflecting the greater exposure of the gut-associated immune system to the enteric parasite. We found that both antibody levels and the parasite burden decreased in repeat infections. This suggests that, possibly due to differences in antibody affinity, only lower antibody levels were required after the development of a sterile immunity after children had been repeatedly exposed to the parasite (Li et al. 2023; Ssewanyana et al. 2017).

The current study had a few limitations to consider. One of them was the challenge of unequivocally linking a diarrhea episode to *Cryptosporidium*, as children in this community were concurrently infected with various enteropathogens (Taniuchi et al. 2013). For most enteric pathogens, at our study site the incidence of diarrhea is highest between 6 months and 3 years of age (Liu et al. 2016; Platts-Mills et al. 2015). To rectify the attribution issue, during this time period Cryptosporidium-associated diarrheal infections were defined as an episode of diarrhea that came with a new Cryptosporidium infection, indicated by the preceding stool sample(s), either from regular surveillance or from a diarrheal episode, testing negative for Cryptosporidium for at least 65 days prior. (Steiner et al. 2018). In years 4 and 5 the study surveillance was decreased in Sub-Group 2 samples which may have resulted in an overestimation of the amount of diarrheal cryptosporidiosis which may therefore decline even more rapidly than is suggested in Figure 2 – to investigate if this was the case the diarrheal data from the children in each group was analyzed separately (Supplemental Figure 1). No difference in the diarrheal infection profile in the two Sub-Groups was observed.

The study did have significant strengths. These included its longitudinal design, which entailed a combination of routine and clinical specimen collection, complemented by qPCR and ELISA testing. A vaccine delivered prior to the rainy season when the risk of cryptosporidiosis is high could significantly reduce post-neonatal cryptosporidiosis and improve child health by reducing the burden of diarrheal disease (Gilchrist et al. 2018; Wutor et al. 2023).

## Methods

### Child cohort

500 children were enrolled before 1 week of birth within an impoverished neighborhood of Dhaka, Bangladesh between June 2014 and March 2016 (Figure 1). These children were monitored for diarrhea through bi-weekly visits to their home by trained field investigators (“Cryptosporidiosis and Enteropathogens in Bangladesh”; ClinicalTrials.gov identifier NCT02764918). Fecal samples collected from both all diarrheal episodes and asymptomatic surveillance were processed to extract fecal DNA to run a diagnostic qPCR assay to detect the protozoan parasites *Cryptosporidium spp*., *Giardia lamblia* and *Entamoeba histolytica* (Steiner et al. 2018). At our Bangladesh study site, disease caused by the protozoan parasites was expected to be common, and a particular cause of concern during the post-neonatal period (Black et al. 2013; Haque et al. 2009; Korpe et al. 2016; Mondal et al. 2012; Steiner et al. 2018). However, in 2019 when most of the children were close to completing the third year of study the protocol was modified to extend the follow-up beyond 3 years. This resulted in a) Sub-Group1 of children with uninterrupted bi-weekly diarrheal surveillance and monthly asymptomatic stool collection and b) Sub-Group2 with collection of diarrheal samples available during clinic visit during the episode and bi-annual asymptomatic stool collection. To study the potential role of mucosal and systemic immunity, 54 children were randomly selected from among the children who had at least one cryptosporidium infection prior to 1 year of age (Figure 1, 4).

In accordance with WHO polio eradication protocols, stool samples collected prior to May 2016 were discarded after molecular diagnostic assays were performed to identify symptomatic and asymptomatic infections these were not therefore available for fecal IgA analysis (Poliovirus Containment Advisory Group 2022). Plasma samples were also collected from the children bi-annually for later antibody level analysis.

### Human Sampling and Specimen Testing

Stool samples collected fresh in the field were placed in ice and deposited into the icddr,b laboratory by the end of the same day, where they were subsequently frozen at -80°C within 6 hours of collection. Blood samples collected at the Study Clinic were brought to the lab biobank for processing, freezing and storage. A subset of fifty-four children (Figure 1, 4) who were identified as infected with the Cryptosporidium parasite prior to one year in age were selected to investigate the anti-Cp23 and Cp27 IgA and IgG immune profile using the fecal samples collected after May 2016 and the plasma samples. Blood samples were collected biannually from the children during years 1-3 and annually at years 4 and 5 of age (Figure 2B).

### ELISA Protocol to detect anti-Cp23 and Cp27 IgG and IgA antibodies

Fecal dilution buffer was first prepared consisting of PBS mixed with 0.5M EDTA pH 8, 50μg/ml of soybean trypsin inhibitor, and 666μL of 100mM PMSF. Fecal samples were then thawed and a single 1100 mg aliquot of stool was combined 1:10 in Fecal Dilution Buffer. One 5mL Eppendorf capful of glass beads was added, and the sample was homogenized in a Qiagen TissueLyser II for 10 minutes. Samples were then centrifuged at 10,000 g for 5 minutes. Supernatants were collected and stored for ELISA antibody testing.

ELISA plates were coated with 50μL per well of the CP17 or CP23 antigen solution at a concentration of 5 μg/mL, diluted in PBS. The plate was covered and left to incubate overnight on a shaker at 4 degrees. The plate was then washed 3 times with PBS mixed with 0.05% Tween 20 (PBST) wash buffer to remove any non-adhered coating buffer. 150 μL of ELISA blocking solution was added to each well, and the plate was allowed to incubate for 1 hour at 37 degrees in a shaking incubator. The wells were then washed one more time. 50 μL of fecal sample diluted 1:4 in blocking buffer were added to each well on the plate. The plate was again incubated for an hour at 37 degrees in the shaking incubator. The plate was then washed 3 times in PBST wash buffer to remove the primary fecal sample. 50 μL of HRP antihuman conjugated secondary antibody (Southern Biotechnology) was added per well, and left to incubate for 1 hour at 37 degrees. The plate was then washed 3 times with PBST and 50 μL per well of TMB Turbo ELISA Substrate (Fisher ThermoScientific) was added. The plates were incubated at room temperature (RT) for 7.5 minutes, then 50 μL of stop solution (2N H2SO4) was added to each well. Absorbance was measured at 450 nm (corrected with absorbance at 570 nm). For plasma samples, the same procedure was carried out except that the dilution of the primary samples was 1500x for IgG and 150x for IgA. In all plates, a dilution series of a pool of strong positive samples was used to generate a standard curve and determine the relative antibody values, and negative control samples were used to determine a positive cutoff value.

### Statistical Analysis

Analysis of infection frequency and the impact of repeat infections on parasite burden and antibody levels was performed using the Prism 10 computer program using either the Brown and Wilson Chi-square test for trend or the Kruskai-Wallis test with the Dunn’s test for multiple comparisons of nonparametric data as appropriate (GraphPad) (Brown et al. 2001; McKight & Najab 2010). All other analyses were performed using R (version 4.3.0), with packages tidyverse (version 2.0.0) and readxl (version 1.4.2) used for data organization and visualization (RStudio Team 2015). The strength of the antibody associations with time was analyzed using Pearson’s correlation test and linear regression analysis. All code used for these analyses will be made public with the publication of the paper through GitHub.

### Study approval

The study was approved by the Ethical and Research Review Committees of the International Centre for Diarrhoeal Disease Research, Bangladesh (PR-13092) and the Institutional Review Board of the University of Virginia (IRB# 20388). The ClinicalTrials.gov identifier is NCT02764918. Informed written consent was obtained from the parents or guardians for the participation of their child in the study.

## Data Availability

Data will be deposited in the University of Virginia institutional repository LibraData

https://doi.org/10.18130/V3/ZBILE6

## Acknowledgments

We wish to thank the field workers, nurses, laboratory staff of the Parasitology Laboratory of icddr,b who worked for this project, and parents and children at the icddr,b study sites who participated in this study and without whom we could not have completed this research. We also want to thank our colleagues Farhad Hossain and Sultan Mahmud for their assistance and advice during this project. Work at icddr,b is supported by their core donors (Government of the People’s Republic of Bangladesh, GAC, Sida, and UKAid).

## Author contributions

Drafting of the manuscript was performed by WAOP and CAG. All authors edited and approved the final manuscript. CAG, WAP and RH conceived of the analysis plan and WAOP and BH performed the experiments. WAP and RH founded the birth cohort and directed the study. UN curated and maintained the study database. Data analysis was performed by HHS and GBM. Fieldwork and data collection at the International Centre for Diarrhoeal Disease Research, Bangladesh (icddr,b) were performed by MA and MK, with supervision from ZN, ASGF and RH.

## Supplemental Figure

**Supplemental Figure 1:**
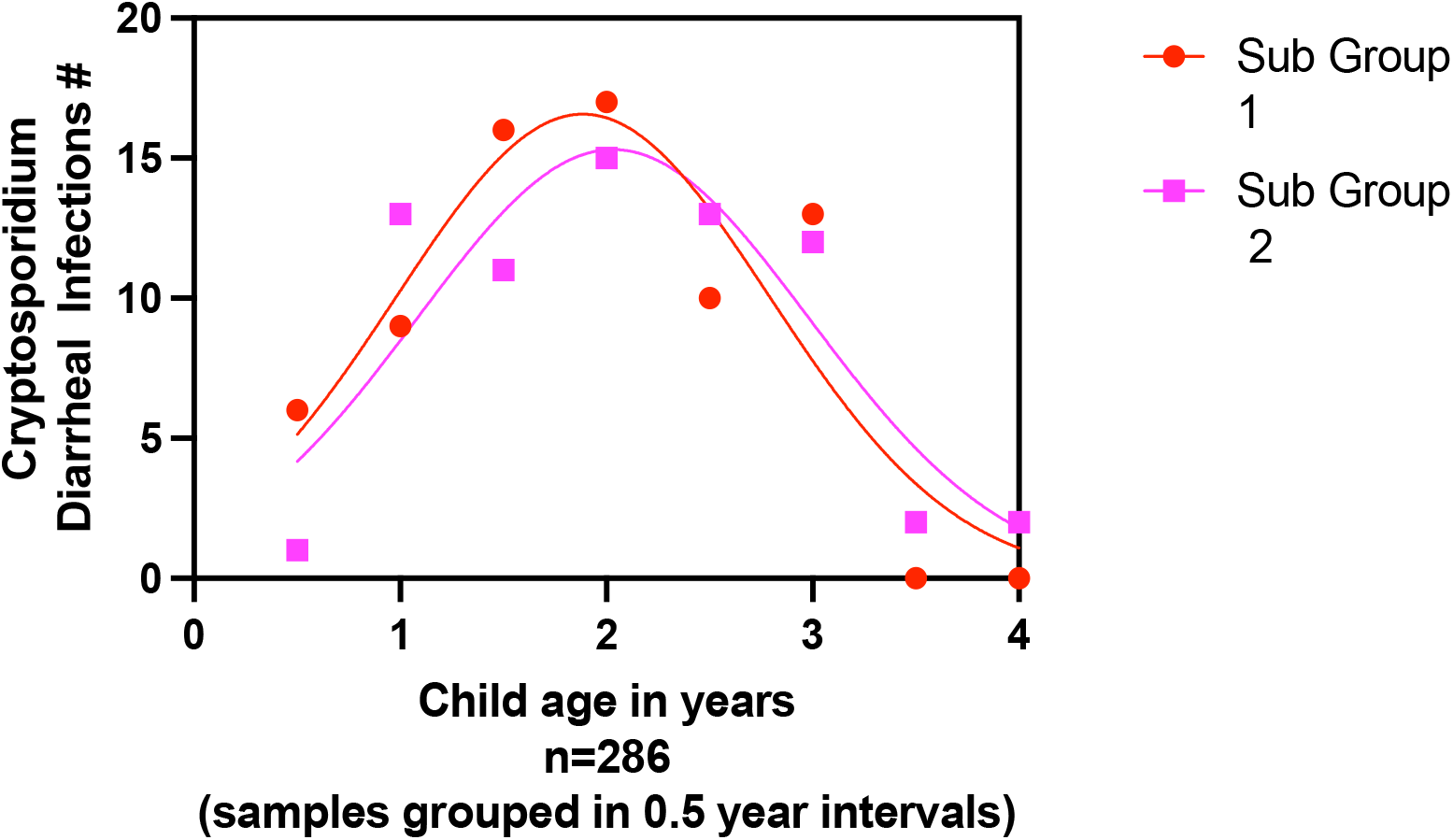
No difference in the prevalence of Diarrheal Cryptosporidiosis was observed when surveillance of asymptomatic disease was decreased in Sub-Group 2. Data shown is only from the 286 children who remained in the study for the entire 5 years. After year 3 children were split into the 2 groups. Sub-Group1. (red symbols and line) children with uninterrupted bi-weekly diarrheal surveillance and monthly asymptomatic stool collection and Sub-Group2 (purple symbols and line) children where diarrheal sample continued to be collected but only bi-annual asymptomatic stools were collected. Infections occurring within each 6-month interval were grouped and the number shown on the Y axis. X axis indicates child age in years

**Supplemental Figure 2.**
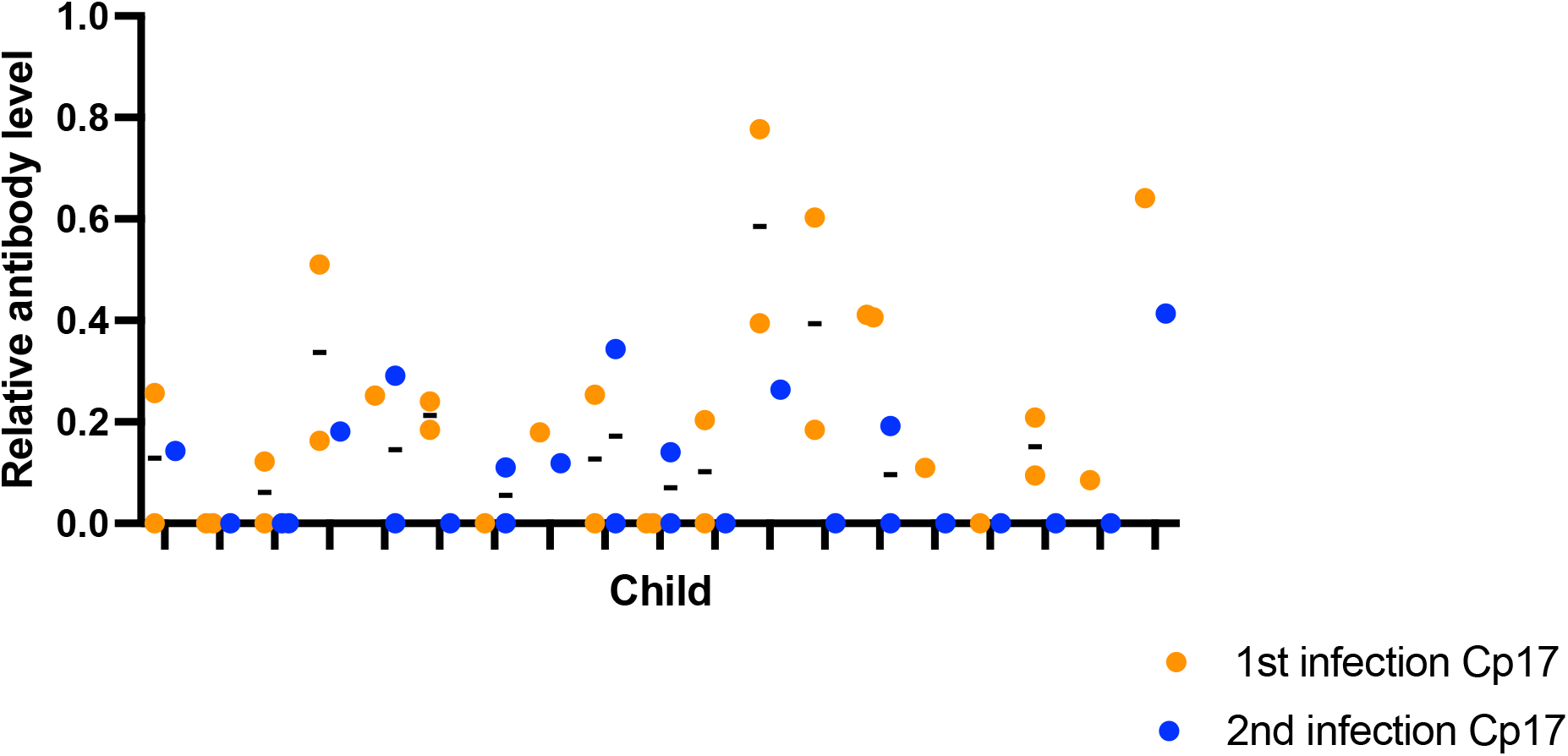
Parasite burden and anti-Cp17 fecal IgA decline in repeat infections. Analysis was restricted to 1^st^ and 2^nd^ infections and to children in which fecal IgA was able to be measured during both infections and the sample had a Cryptosporidium positive Cq value. The number of repeat *Cryptosporidium* infections per child was defined using the entire data set as previously defined (Kabir et al 2021). X value indicates individual children, Y value the relative anti-Cp17 antibody levels (n=19). Values from samples collected during the same infection were included as repeated measurements. Orange symbols indicate samples collected during the first infection, blue symbols indicate samples collected during the second infection. Analysis was done using the mixed model. The difference in the mean antibody levels was 0.11 ± 0.0344 and was statistically significant p=0.0041

**Supplemental Table 1.**
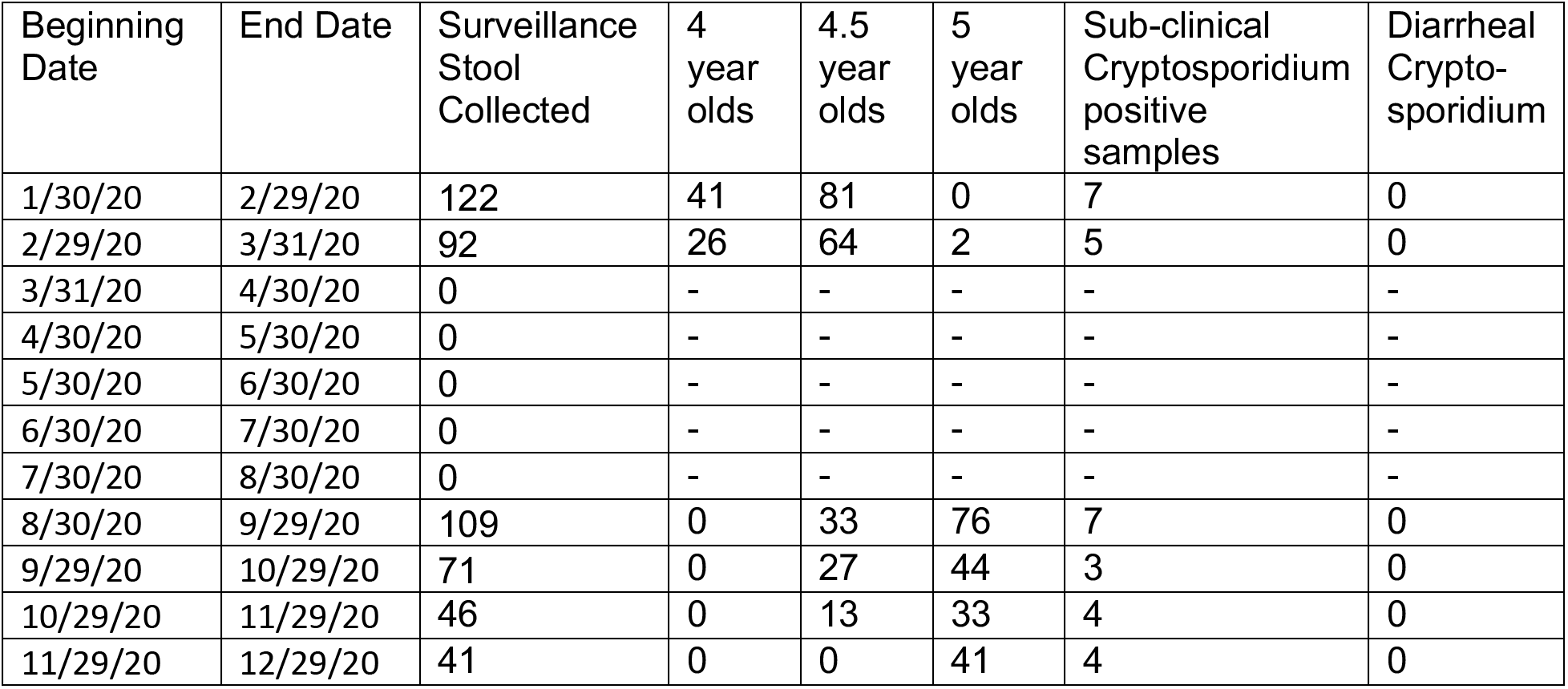
Surveillance Stool Collection During 2020.

**Supplemental Table 2.**
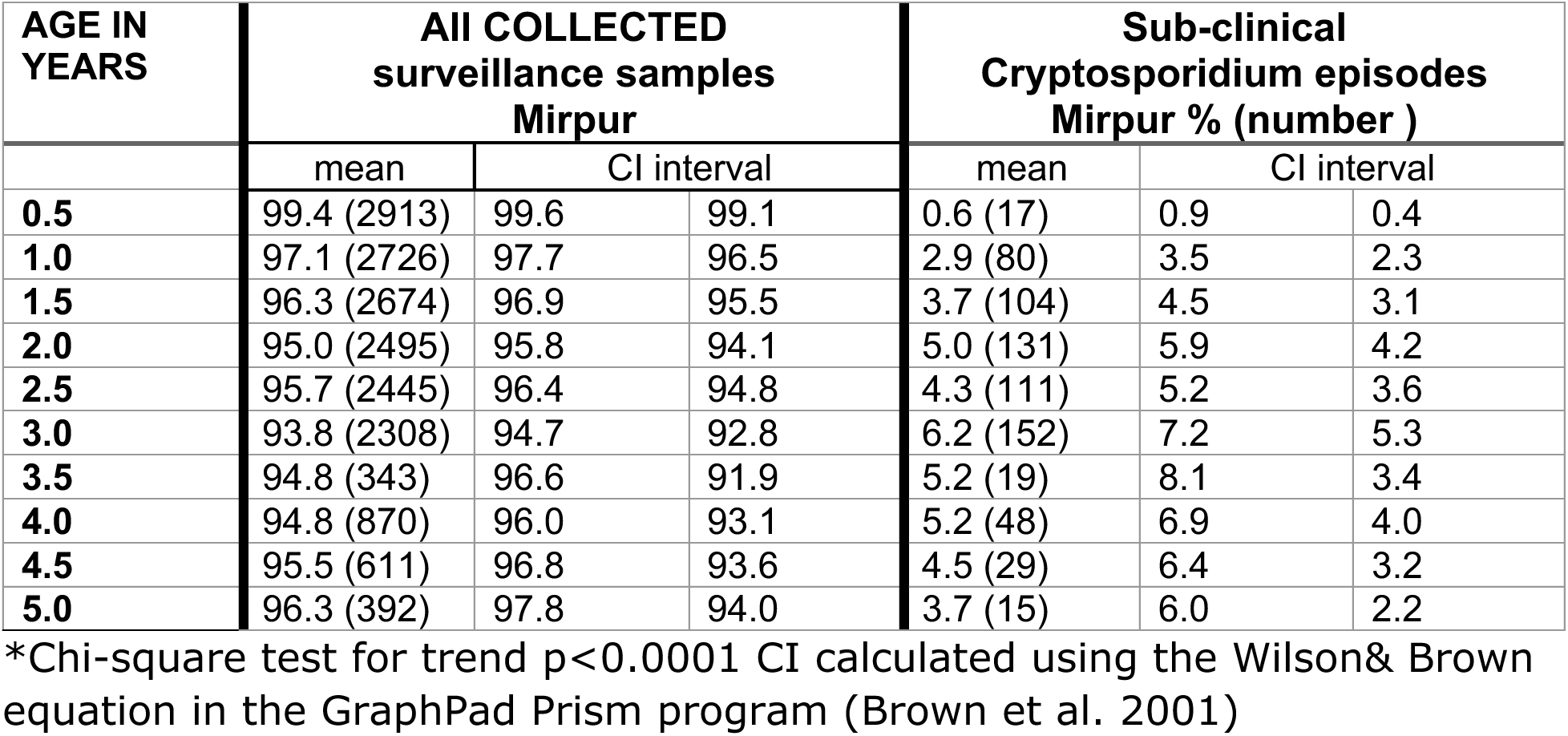
Surveillance Stool collection.

| AGE IN YEARS | All COLLECTED surveillance samples Mirpur |  |  | Sub-clinical Cryptosporidium episodes Mirpur % (number ) |  |  |
| --- | --- | --- | --- | --- | --- | --- |
|  | mean | CI interval |  | mean | CI interval |  |
| **0.5** | 99.4 (2913) | 99.6 | 99.1 | 0.6 (17) | 0.9 | 0.4 |
| **1.0** | 97.1 (2726) | 97.7 | 96.5 | 2.9 (80) | 3.5 | 2.3 |
| **1.5** | 96.3 (2674) | 96.9 | 95.5 | 3.7 (104) | 4.5 | 3.1 |
| **2.0** | 95.0 (2495) | 95.8 | 94.1 | 5.0 (131) | 5.9 | 4.2 |
| **2.5** | 95.7 (2445) | 96.4 | 94.8 | 4.3 (111) | 5.2 | 3.6 |
| **3.0** | 93.8 (2308) | 94.7 | 92.8 | 6.2 (152) | 7.2 | 5.3 |
| **3.5** | 94.8 (343) | 96.6 | 91.9 | 5.2 (19) | 8.1 | 3.4 |
| **4.0** | 94.8 (870) | 96.0 | 93.1 | 5.2 (48) | 6.9 | 4.0 |
| **4.5** | 95.5 (611) | 96.8 | 93.6 | 4.5 (29) | 6.4 | 3.2 |
| **5.0** | 96.3 (392) | 97.8 | 94.0 | 3.7 (15) | 6.0 | 2.2 |

\*Chi-square test for trend $p < 0.0001$ CI calculated using the Wilson & Brown equation in the GraphPad Prism program (Brown et al. 2001)

